# Five central adiposity-based anthropometric indices and cognitive impairment in elderly populations: Development and validation of a risk prediction nomogram using NHANES and CHARLS cohort

**DOI:** 10.1101/2025.07.03.25330791

**Authors:** Jian Chen, Jinzhi Guan, Xuanyu Lu, Yanxiang Liu, Leyi Zhang, Yaxuan Zhang, Shengbing Xue, Jingling Chang

## Abstract

**Background:** Cognitive impairment is a primary contributor to disability among older adults, with growing evidence identifying central adiposity as an adjustable risk factor for neurodegeneration. This study aimed to establish and verify a model for predicting mild cognitive impairment (MCI) risk in aging individuals by incorporating central adiposity indices.

**Methods:** We calculated five central adiposity indices from anthropometric measurements using data from 2,464 United States adults aged ≥ 60 (National Health and Nutrition Examination Survey 2011–2014). Cognitive performance was assessed using three standardized neuropsychological tests. Multivariable logistic regression, incorporating restricted cubic splines to test for nonlinearity, was used to evaluate associations between central obesity measures and MCI, complemented by sensitivity analyses (subgroup stratification and multiple imputation). A random assignment placed participants into either a training (n = 1725) or a internal validation (n = 739) set. Furthermore, the data from participants in the 2011 wave of the China Health and Retirement Longitudinal Study served as an external validation cohort (n = 536). Least absolute shrinkage and selection operator-selected predictors were employed to inform multivariable logistic regression modeling. Assessment of the nomogram’s performance involved the area under the curve (AUC), calibration curve, decision curve analysis, clinical impact curve, and external validation.

**Results:** Positive linear relationships were found between three anthropometric indices of central obesity—a body shape index (ABSI), conicity index (CoI) and weight-adjusted-waist index (WWI)—with MCI risk (*P* < 0.05, *P* for nonlinearity > 0.05). Ten features were screened as MCI predictors, including ABSI, age, educational level, race, poverty income ratio, stroke, depression, diabetes mellitus, physical activity, and gender. The nomogram demonstrated strong discriminative capacity (training AUC = 0.861; internal validation AUC = 0.826; external validation AUC = 0.798), precise calibration, and good clinical utility.

**Conclusion:** The risk of MCI was independently linked to central adiposity indices (ABSI, WWI, and CoI). The nomogram incorporating ABSI provided a validated, clinically applicable prediction model for initial screening of MCI in older populations.

## Introduction

Cognitive dysfunction is a prevalent health concern in aging populations, manifesting as difficulties in memory retention, information processing, sustained attention, and executive functioning. With accelerating global demographic aging trends, age-related cognitive deterioration is projected to increase significantly [1]. Epidemiological meta-analyses have demonstrated that mild cognitive impairment (MCI) affects over 15% of adults aged ≥ 50 years who live in the community [2]. MCI represents an intermediate prodromal stage bridging typical aging and dementia onset, serving as a critical window preceding accelerated cognitive deterioration [3]. According to current estimates, the number of dementia cases around the world is expected to rise sharply, with an anticipated increase of 95.4 million from 2019 to 2050 [4]. These cognitive disorders substantially burden affected individuals, caregivers, and healthcare systems [5]. Despite growing needs, validated screening instruments for early detection of cognitive impairment remain scarce, particularly in resource-limited rural and community healthcare environments [6–8]. Therefore, recognizing practical and accessible risks or protective factors associated with cognition is crucial for the prompt diagnosis and management of cognitive dysfunction.

Obesity constitutes a significant public health crisis in modern society, affecting 71.2% of adults in the United States (U.S.) [9]. Excess visceral fat, particularly central adiposity, has been implicated in cognitive dysfunction [10]. Central obesity increases dementia risk by 28% overall and by 39% among women compared to non-centrally obese counterparts [11]. Despite its widespread use for classifying obesity, the limitations of body mass index (BMI) are becoming more evident. An analysis of 29 long-term cohort studies (n = 20,083) indicated that elevated BMI during midlife is linked to a higher risk of dementia, whereas an elevated BMI in late life was associated with reduced risk, reflecting the debated “obesity paradox” [12,13]. Critically, BMI neither distinguishes fat from muscle mass nor reflects regional fat deposition [14], and individuals with a normal BMI may still exhibit aberrant adiposity [15]. Emerging indices of anthropometry—including a body shape index (ABSI), weight-adjusted-waist index (WWI), body roundness index (BRI), conicity index (CoI), and waist-to-height ratio (WHtR)—derived from waist circumference (WC), body height (BH), and body weight (BW), offer superior assessment of central obesity compared to BMI [16,17]. While prior studies have linked select central adiposity-based anthropometric indices to cognitive functioning [18–20], comprehensive predictive models integrating these measures for cognitive decline in aging populations remain underexplored.

In light of these findings, this study aims to systematically assess the relationships between five abdominal adiposity anthropometric indices and cognitive dysfunction among U.S. adults aged 60 and older. Furthermore, it seeks to develop a straightforward screening instrument with significant predictive validity to identify seniors at-risk early, thereby facilitating targeted preventive measures and clinical interventions.

## Materials and methods

### Data source and study participants

The National Health and Nutrition Examination Survey (NHANES), administered by the National Center for Health Statistics (NCHS) as part of its national surveillance system, systematically monitors population health and nutritional status through multimodal data collection. This nationally representative program was used to collect comprehensive datasets encompassing demographic profiles, biochemical measurements, and health-related questionnaires. Following NCHS Institutional Review Board approval and documented participant consent, we analyzed pooled data from the 2011 to 2014 survey cycles, and all participants provided written informed consent, initially comprising 19,931 respondents [21]. NHANES was conducted in accordance with the ethical principles of the Declaration of Helsinki. The study utilized publicly available data that was de-identified. Cognitive assessments were restricted to participants aged ≥ 60 years (n = 3,632). After applying exclusion criteria for missing (a) cognitive test results (n = 698), (b) anthropometric measurements (height/weight/WC; n = 182), and (c) covariate data (n = 288), the final analytical cohort contained 2,464 eligible individuals (Fig. 1).

**Fig. 1.**
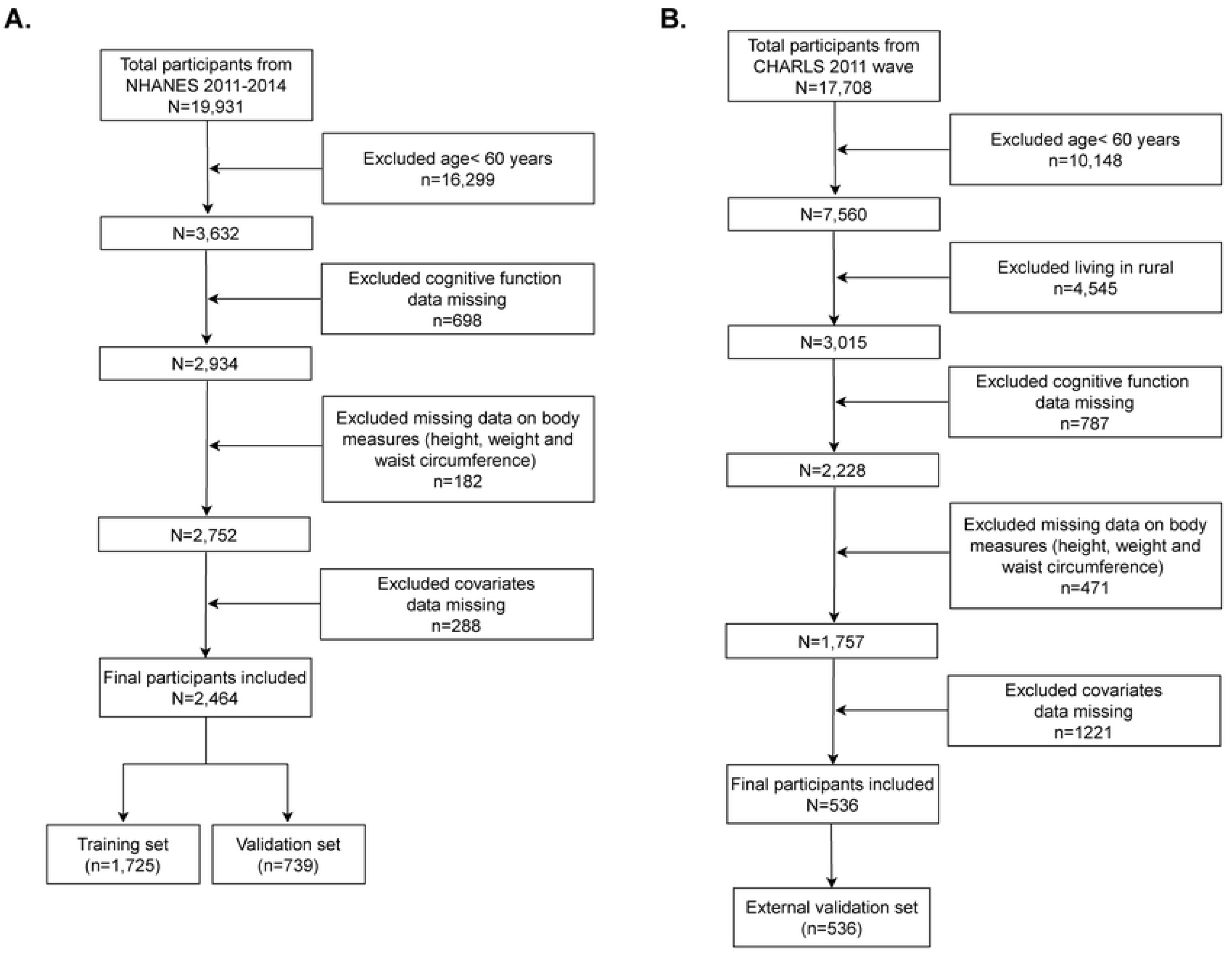
Flowchart of participant selection from the NHANES 2011 to 2014.

### Assessment of cognitive functioning

The NHANES 2011–2014 dataset incorporates three standardized cognitive assessments: the Consortium to Establish a Registry for Alzheimer’s Disease (CERAD) test, the digit symbol substitution test (DSST), and the animal fluency test (AFT) [22–24]. The CERAD test evaluates verbal information encoding through immediate and delayed recall components, featuring three consecutive learning trials followed by a delayed recall trial. Each trial is scored 0–10, with the summed results yielding an immediate recall total score (range: 0–30). The delayed recall total scores (0–10) quantify retention after AFT and DSST completion.

AFT evaluates verbal fluency and executive function through timed semantic categorization. Participants receive one point per unique animal named within 60 s, with cumulative scores reflecting lexical access efficiency [25]. DSST quantifies cognitive processing speed, working memory, and visual attention. During this 120-s task, participants pair symbols with corresponding numbers across 133 items, scoring one point per accurate pairing [26].

Domain-specific Z-scores from individual neuropsychological assessments were standardized and aggregated to derive a composite metric reflecting global cognitive performance. Clinical validity was established by operationalizing psychometric MCI (p-MCI) as composite z-scores falling ≥ 1 standard deviation (SD) below the average of the population [27].

### Calculation of five central adiposity-based anthropometric indices

Certified medical technicians obtained standardized anthropometric measurements, including BW, BH, and WC. Subsequent calculations for ABSI, BRI, CoI, WWI, and WHtR were conducted using the established formulas outlined below:

ABSI was calculated as shown below [28]:

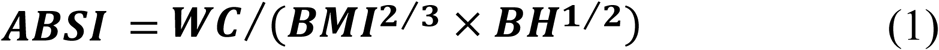

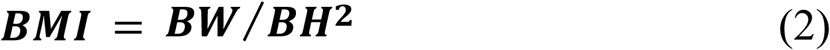

BRI was calculated as shown below [15]:

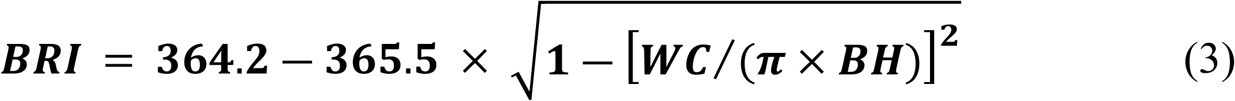

CoI was calculated as shown below [29]:

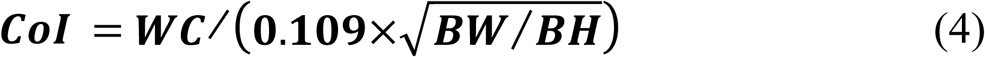

WWI was calculated as shown below [30]:

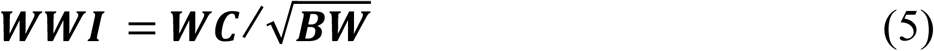

WHtR was calculated as shown below:

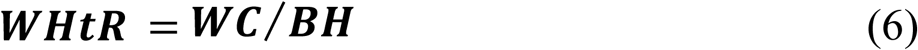

### Assessment of covariates

Through an in-depth review of epidemiological literature and clinical insights, potential covariates connected to sociodemographic factors, lifestyle behaviors, anthropometric indicators, and clinical comorbidities were identified [19,31]. Regarding sociodemographic factors: age cohorts were stratified into 60–69, 70–79, and ≥ 80 years [24]. Racial classification comprised non-Hispanic White, non-Hispanic Black, other Hispanic, Mexican American, and other races. Marital status groupings included never married, widowed/divorced/separated, and married/living with a partner. The poverty-income ratio (PIR) thresholds were > 3.50, 1.31–3.50, and ≤ 1.3 [32]. Education level was tied to above high school, high school or its equivalent, and below high school [33].

Concerning lifestyle behaviors: smoking status was determined through two validated questions: lifetime consumption of ≥ 100 cigarettes and current usage, yielding current, former, and never smoker classifications. Drinking status was classified as current drinkers (subclassified by intake severity), former (12 or more annual drinks without recent consumption), or never (fewer than 12 lifetime drinks) [34]. The physical activity (PA) levels were quantified via weekly metabolic equivalent (MET)– minutes from walking, cycling, work, and recreational activities: sufficiently active (≥ 600), insufficiently active (1–599), and inactive (0) [35].

In relation to clinical comorbidities: hypertension criteria included systolic blood pressure (BP) ≥ 140 mmHg, diastolic BP ≥ 90 mmHg, taking antihypertensive medication, or self-reported diagnosis [36]. Self-reported medical histories confirmed coronary heart disease (CHD), stroke, and diabetes mellitus (DM) status [37]. Depression severity was evaluated with the Patient Health Questionnaire, with clinical depression defined as a score ≥ 10 points [38].

### External validation

The China Health and Retirement Longitudinal Study (CHARLS) tracks individuals aged 45 and older and is representative of the population in China [39]. Peking University’s Institutional Review Board approved the CHARLS protocol. Written consent was secured from all participants before they were enrolled. The study employed publicly accessible data that was de-identified. The data from CHARLS survey wave one, conducted in 2011, were utilized for external validation (n= 17,708). We implemented the following exclusion criteria to ensure the data’s integrity and its appropriateness for external validation: (a) age <60 years or with age data missing (n = 10,148), (b) lived in rural (n= 4,545), (c) cognitive function information missing (n=787), (d) height/weight/WC data missing ( n = 471), (f) covariate data missing (n = 1,221). Ultimately, 536 eligible older adults were included (Fig. 1). Detailed descriptions and assessments of the variables employed in this study were provided in S1 table.

### Statistical analysis

Parametric data were summarized using mean ± SD, while non-parametric variables were reported as median with interquartile range. The representation of categorical measures was done through frequencies and percentages (n%). Comparisons between Groups for continuous measures were conducted using independent *t*-tests for parametric distributions and Mann–Whitney *U* tests for non-parametric distributions. Categorical variable analyses utilized Pearson’s chi-square test where statistically appropriate.

Multivariable logistic regression analyses were constructed to assess covariate-adjusted associations between central obesity indicators and p-MCI, with outcomes expressed as adjusted odds ratios and 95% confidence intervals (CIs). Each index was modeled as continuous variables standardized via Z-scoring (calculated as [observed value - mean]/SD). Covariate selection followed evidence-based criteria integrating prior research and clinical relevance [26,31,40]. Multicollinearity was evaluated using generalized variance inflation factors (GVIF), with GVIF^(1/(2 × degrees of freedom [Df])) ≥ 2 indicating collinearity [41]. Three hierarchical models were established: Model 1 (unmodified); Model 2 (adjusted for age, gender, and race); Model 3 (A fully adjusted model that incorporated all the potential confounding variables identified above). A restricted cubic spline (RCS) with four knots was employed to investigate the potential nonlinear association between each anthropometric index and p-MCI.

Sensitivity analyses included subgroup and multiple imputation analyses. Subgroup analyses were conducted across demographic and clinical categories: age (60–69, 70–79, ≥ 80 years), gender (male and female), racial groups (Non-Hispanic Black, Non-Hispanic White, and other races), PIR (≤ 1.3, > 1.3), education level (above high school, high school or equivalent, and below high school), PA (inactive, insufficiently active, and sufficiently active), drinking status (current, former, never), BMI (≥ 30; 25–30; < 25), and comorbidity status (hypertension/stroke/DM/depression: present/absent). Stratified regression models were used to assess heterogeneity, with interaction effects quantified via likelihood ratio testing. To address missing covariate data (n = 288), multivariate imputation via chained equations was performed to generate five complete datasets using variables from the final analytical framework [34].

The dataset was randomly partitioned into training (n = 1,725) and validation (n = 739) cohorts, employing a 7:3 split ratio. To identify p-MCI predictors, the least absolute shrinkage and selection operator (LASSO) regression analysis was conducted on the training cohort, optimizing λ determination through 10-fold cross-validation [42]. Model selection adhered to the one-standard-error (SE) rule, prioritizing parsimony while retaining predictive accuracy [43]. Candidate predictors were entered into multivariate logistic regression for further refinement to identify independent factors significantly associated with p-MCI. A final multivariable model was constructed using retained predictors, from which a clinical nomogram was developed to graphically represent predictor contributions. This scoring tool quantifies individual risk factors through weighted scoring, where a summative scoring system estimates composite p-MCI probability.

Model performance was comprehensively evaluated across four domains: Predictive accuracy (discrimination), reliability (calibration), clinical utility, and generalizability. The area under the curve (AUC) of the receiver operating characteristic (ROC) curves was employed to measure predictive capacity. Calibration fidelity was verified through two complementary approaches: goodness-of-fit assessment via Hosmer–Lemeshow testing [44] and smoothed calibration curves employing local regression (loess) algorithms to visualize prediction-actual probability alignment. Clinical relevance was assessed using decision curve analysis (DCA) to quantify net intervention benefits [45] and clinical impact curve (CIC) to evaluate risk stratification practicality [46]. Model generalizability was evaluated through internal and external cohort performance metrics.

All analytical workflows were implemented using R software (version 4.3.1; R Foundation) and FreeStatistics (version 2.1.1; FreeClinical Medical Technology). Statistical significance thresholds were set at α = 0.05 (two-tailed).

## Results

### Baseline characteristics of the participants

The analytical cohort comprised 2,464 eligible participants from NHANES 2011 to 2014, including 418 individuals meeting p-MCI diagnostic criteria. Comparative analyses revealed significant between-group differences (*P* < 0.05) in sociodemographic variables (age, gender, race, marital status, education level, and PIR), lifestyle factors (drinking status and PA), clinical comorbidities (hypertension, stroke, DM, and depression), and central adiposity indices (ABSI, WWI, and CoI). Non-significant intergroup variations (*P* > 0.05) were observed for smoking status, CHD, BMI, WC, BRI, and WHtR (Table 1).

**Table 1.** Baseline characteristics of the participants.

| Characteristic | Overall<br>(*N* = 2464) | Non p-MCI<br>(*N* = 2046) | p-MCI<br>(*N* = 418) | *P*-value |
| --- | --- | --- | --- | --- |
| **Age, years, n (%)** |  |  |  | < 0.001 |
| 60–69 | 1368 (55.5) | 1208 (59) | 160 (38.3) |  |
| 70–79 | 726 (29.5) | 572 (28) | 154 (36.8) |  |
| ≥ 80 | 370 (15.0) | 266 (13) | 104 (24.9) |  |
| **Gender, n (%)** |  |  |  | 0.005 |
| Male | 1196 (48.5) | 967 (47.3) | 229 (54.8) |  |
| Female | 1268 (51.5) | 1079 (52.7) | 189 (45.2) |  |
| **Race, n (%)** |  |  |  | < 0.001 |
| Non-Hispanic White | 1199 (48.7) | 1080 (52.8) | 119 (28.5) |  |
| Non-Hispanic Black | 576 (23.4) | 441 (21.6) | 135 (32.3) |  |
| Mexican American | 212 (8.6) | 158 (7.7) | 54 (12.9) |  |
| Other Hispanic | 248 (10.1) | 169 (8.3) | 79 (18.9) |  |
| Other Races | 229 (9.3) | 198 (9.7) | 31 (7.4) |  |
| **Marital status, n (%)** |  |  |  | < 0.001 |
| Married/living with a partner | 1435 (58.2) | 1235 (60.4) | 200 (47.8) |  |
| Widowed/divorced/separated | 890 (36.1) | 694 (33.9) | 196 (46.9) |  |
| Never married | 139 (5.6) | 117 (5.7) | 22 (5.3) |  |
| **Education level, n (%)** |  |  |  | < 0.001 |
| Below high school | 583 (23.7) | 336 (16.4) | 247 (59.1) |  |
| High school or equivalent | 573 (23.3) | 485 (23.7) | 88 (21.1) |  |
| Above high school | 1308 (53.1) | 1225 (59.9) | 83 (19.9) |  |
| **PIR, n (%)** |  |  |  | < 0.001 |
| ≤ 1.3 | 721 (29.3) | 505 (24.7) | 216 (51.7) |  |
| 1.3–3.5 | 947 (38.4) | 801 (39.1) | 146 (34.9) |  |
| > 3.5 | 796 (32.3) | 740 (36.2) | 56 (13.4) |  |
| **Smoking status, n (%)** |  |  |  | 0.514 |
| Never | 1209 (49.1) | 1008 (49.3) | 201 (48.1) |  |
| Former | 949 (38.5) | 791 (38.7) | 158 (37.8) |  |
| Current | 306 (12.4) | 247 (12.1) | 59 (14.1) |  |
| **Drinking status, n (%)** |  |  |  | < 0.001 |
| Never | 373 (15.1) | 284 (13.9) | 89 (21.3) |  |
| Former | 691 (28.0) | 527 (25.8) | 164 (39.2) |  |
| Current | 1400 (56.8) | 1235 (60.4) | 165 (39.5) |  |
| **PA, n (%)** |  |  |  | < 0.001 |
| Inactive | 813 (33.0) | 627 (30.6) | 186 (44.5) |  |
| Insufficiently active | 421 (17.1) | 347 (17) | 74 (17.7) |  |
| Sufficiently active | 1230 (49.9) | 1072 (52.4) | 158 (37.8) |  |
| **Hypertension, n (%)** |  |  |  | < 0.001 |
| No | 709 (28.8) | 631 (30.8) | 78 (18.7) |  |
| Yes | 1755 (71.2) | 1415 (69.2) | 340 (81.3) |  |
| **Stroke, n (%)** |  |  |  | < 0.001 |
| No | 2301 (93.4) | 1934 (94.5) | 367 (87.8) |  |
| Yes | 163 ( 6.6) | 112 (5.5) | 51 (12.2) |  |
| **CHD, n (%)** |  |  |  | 0.448 |
| No | 2246 (91.2) | 1869 (91.3) | 377 (90.2) |  |
| Yes | 218 (8.8) | 177 (8.7) | 41 (9.8) |  |
| **DM, n (%)** |  |  |  | < 0.001 |
| No | 1895 (76.9) | 1617 (79) | 278 (66.5) |  |
| Yes | 569 (23.1) | 429 (21) | 140 (33.5) |  |
| **Depression, n (%)** |  |  |  | < 0.001 |
| No | 2246 (91.2) | 1889 (92.3) | 357 (85.4) |  |
| Yes | 218 ( 8.8) | 157 (7.7) | 61 (14.6) |  |
| **BMI (kg/m<sup>2</sup>), Mean ± SD** | 29.08 ± 6.24 | 29.17 ± 6.27 | 28.62 ± 6.07 | 0.099 |
| **WC (cm), Mean ± SD** | 102.10 ± 14.73 | 102.19 ± 14.73 | 101.64 ± 14.74 | 0.489 |
| **ABSI, Mean ± SD** | 0.08 ± 0.00 | 0.08 ± 0.00 | 0.09 ± 0.00 | < 0.001 |
| **WWI, Mean ± SD** | 11.49 ± 0.71 | 11.45 ± 0.70 | 11.66 ± 0.74 | < 0.001 |
| **CoI, Mean ± SD** | 1.35 ± 0.08 | 1.35 ± 0.08 | 1.37 ± 0.08 | < 0.001 |
| **BRI, Mean ± SD** | 5.98 ± 2.13 | 5.96 ± 2.12 | 6.10 ± 2.21 | 0.225 |
| **WHtR, Mean ± SD** | 0.62 ± 0.09 | 0.62 ± 0.09 | 0.62 ± 0.09 | 0.24 |
| **IRT, Mean ± SD** | 19.11 ± 4.59 | 20.12 ± 4.01 | 14.16 ± 3.98 | < 0.001 |
| **DRT, Mean ± SD** | 6.02 ± 2.29 | 6.49 ± 2.06 | 3.76 ± 1.99 | < 0.001 |
| **AFT, Mean ± SD** | 16.84 ± 5.50 | 17.89 ± 5.20 | 11.67 ± 3.71 | < 0.001 |
| **DSST, Mean ± SD** | 46.75 ± 16.98 | 51.61 ± 13.94 | 22.97 ± 8.24 | < 0.001 |
| **CF Z-score, Median (IQR)** | 0.0 (−0.7, 0.7) | 0.2 (−0.3, 0.9) | −1.4 (−1.7, −1.2) | < 0.001 |

### Associations between five central adiposity-based anthropometric indices with p-MCI

The multivariable-adjusted associations between the central adiposity indices and p-MCI risk are depicted in Fig. 2. Multicollinearity assessment revealed acceptable variance inflation (GVIF^(1/(2 × Df) < 2 for all covariates), confirming model stability (S2 Table). Three indices—ABSI, WWI, and CoI—demonstrated significant positive associations with p-MCI across all adjustment levels (*P* < 0.05). The RCS analyses confirmed linear relationships for these indices in fully adjusted models (*P* for nonlinearity > 0.05; Fig. 3).

**Fig. 2.**
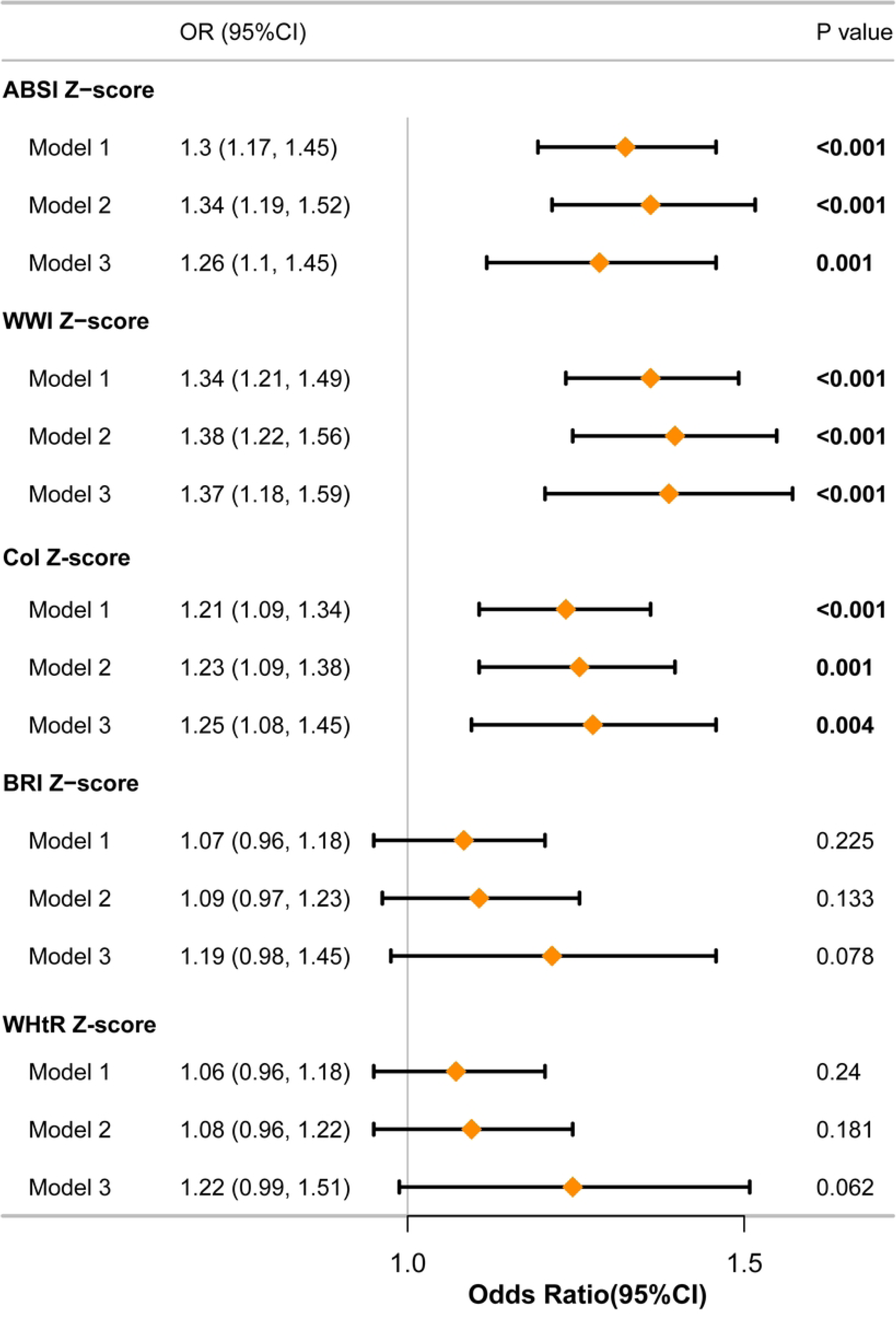
Associations between five central adiposity-based anthropometric indices and p-MCI. Model 1 included no covariate adjustments. Model 2 was controlled for demographic factors (age, gender, and race). Model 3 was additional adjustments for demographic factors (marital status, education level, and PIR), lifestyle (smoking status, drinking status, and PA), anthropometric index (BMI), and clinical comorbidities (hypertension, stroke, CHD, DM, and depression).

**Fig. 3.**
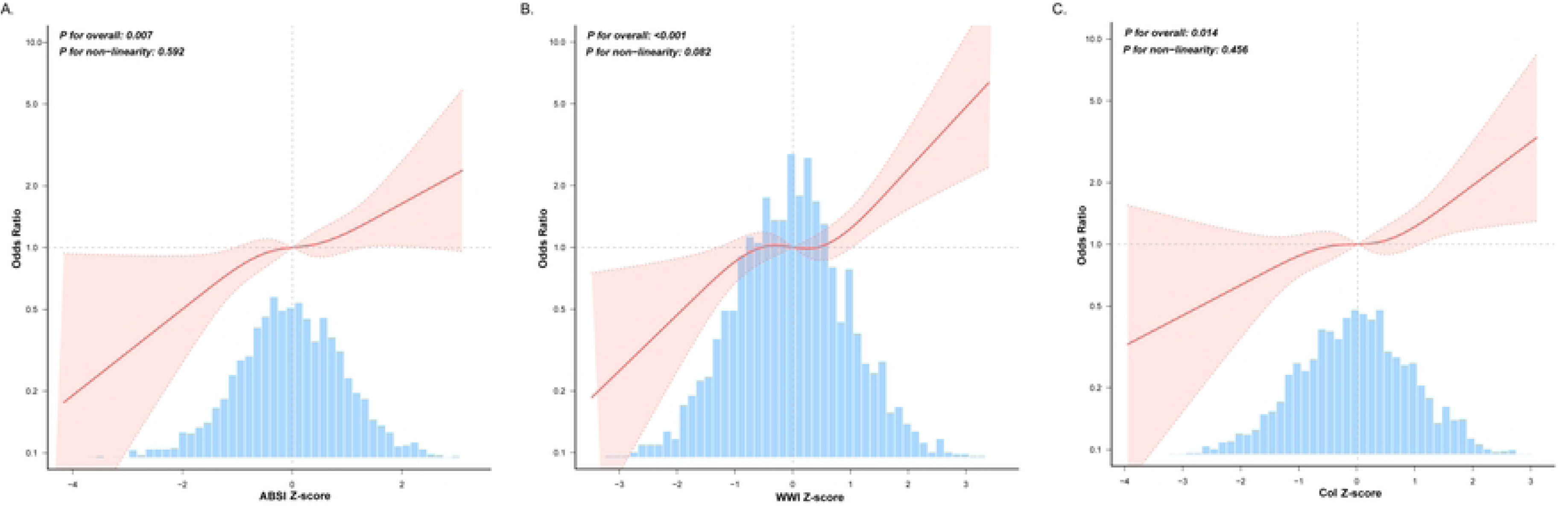
Restricted cubic spline relationships between central adiposity-based anthropometric indices and p-MCI. **(A)** Relationship between ABSI and p-MCI; **(B)** Relationship between WWI and p-MCI; **(C)** Relationship between CoI and p-MCI. Model was adjusted for demographic factors (age, gender, race, marital status, education level, and PIR), lifestyle (smoking status, drinking status, and PA), anthropometric index (BMI), and clinical comorbidities (hypertension, stroke, CHD, DM, and depression).

### Sensitivity analysis

The sensitivity analysis validated the robustness of the primary findings. Stratified analyses evaluated effect modification across various subgroups for the associations of ABSI, WWI, and CoI with p-MCI (S1–S3 Figs.). The ABSI demonstrated consistent positive associations with p-MCI risk, with no significant interaction effects in any subgroup (*P* for interaction > 0.05), confirming its stability across different variables (S1 Fig.). For WWI and CoI, PIR emerged as a significant effect modifier (P for interaction < 0.05; S2 and S3 Figs.). The subgroup analyses demonstrated that ABSI is applicable and robust across various populations, confirming its reliability as a predictor of p-MCI risk.

The multiple imputation analysis yielded consistent results, with the positive associations between ABSI, WWI, and CoI and the risk of p-MCI remaining significant across all three models (*P* < 0.05; S3 Table).

### Predictor selection

Based on the findings from multiple logistic regression analyses, RCS regression analyses, subgroup analyses, and multiple imputation analyses, ABSI, WWI, and CoI were retained for predictor selection. Baseline characteristics of the training (n = 1,725) and internal validation (n = 739) cohorts were comparable across all 18 evaluated parameters (*P* > 0.05; Table 2). The LASSO regression with cross-validation identified 12 candidate predictors from the training cohort (Fig. 4). Subsequent multivariable logistic regression incorporating these predictors revealed 10 statistically significant p-MCI risk factors (*P* < 0.05): ABSI, age, educational level, race, PIR, stroke, depression, DM, PA, and gender (Fig. 5). Furthermore, we used the data from the CHARLS 2011 wave as an external validation cohort. The characteristics of 10 predictors were shown in S4 table.

**Fig. 4.**
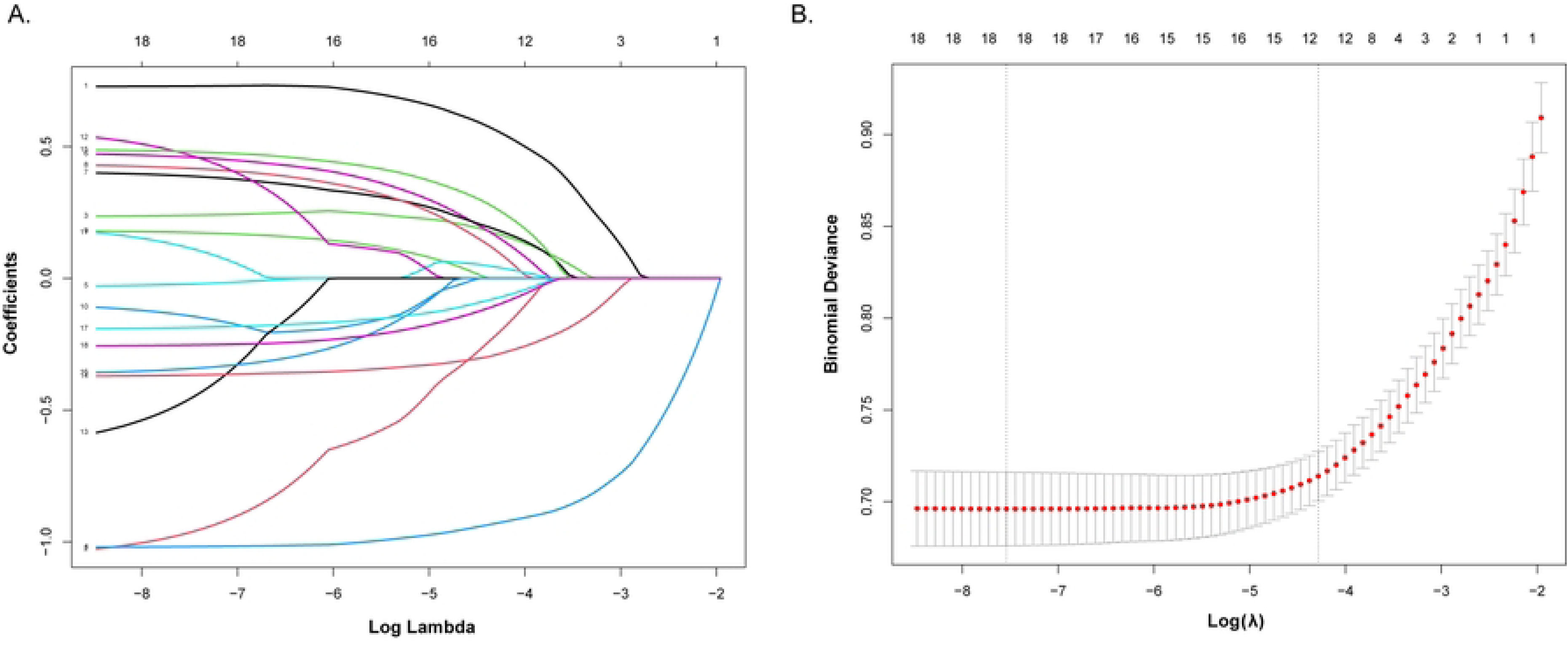
LASSO regression analysis of predictors for p-MCI. **(A)** The regularization path analysis illustrated coefficient trajectories across a logarithmic λ sequence, with the optimal λ value retaining 12 non-zero coefficients. **(B)** Tenfold cross-validation identified the parsimonious λ value using the one SE rule, selecting the simplest model within one SE of the minimum binomial deviance. This approach balances predictive accuracy with model simplicity to mitigate overfitting.

**Fig. 5.**
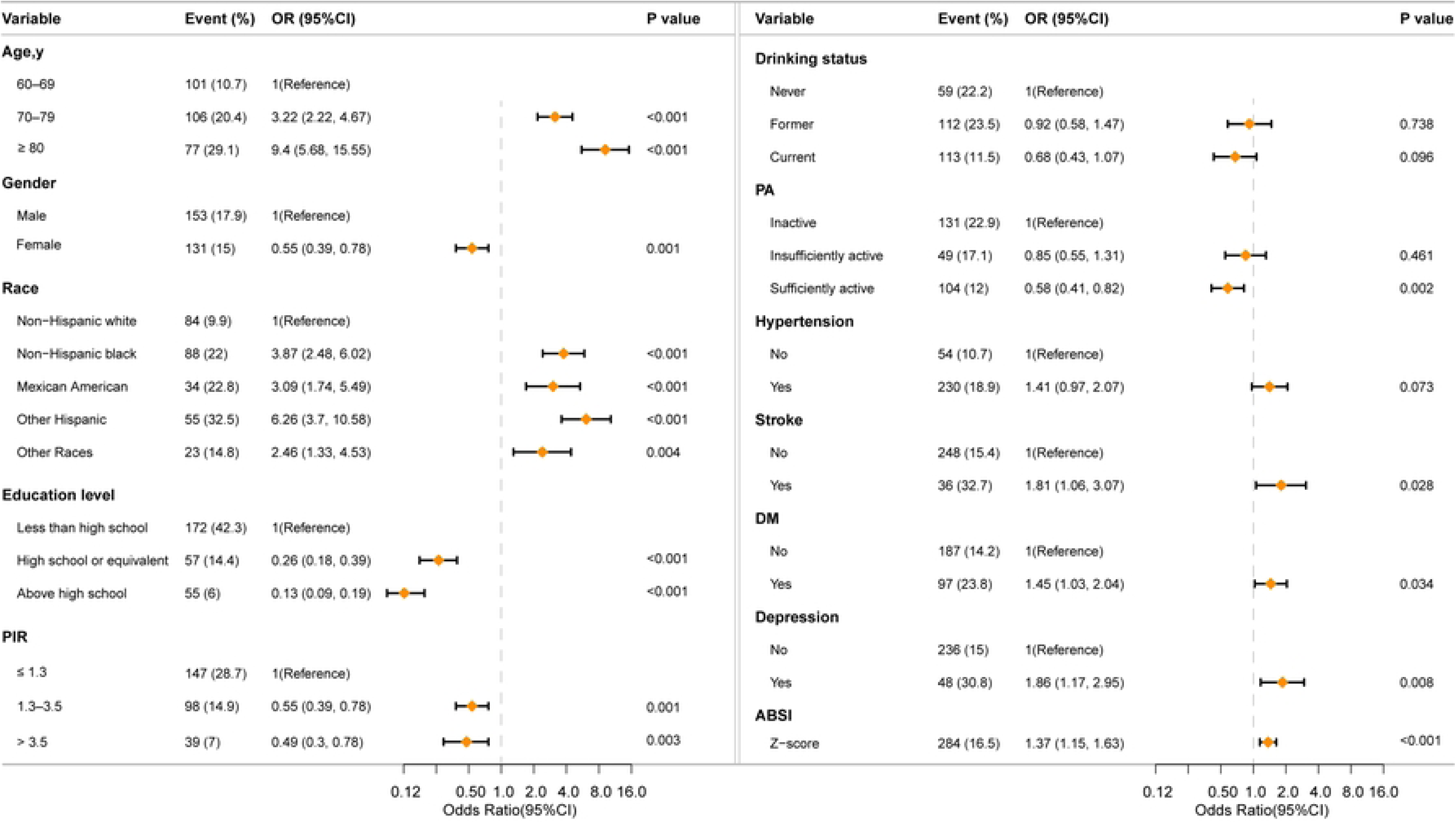
Multivariate logistic regression analysis of predictors for p-MCI.

**Table 2.** Baseline characteristics between training set and validation set.

| Characteristic | Training set<br>(*N* = 1725) | Validation set<br>(*N* = 739) | *P*-value |
| --- | --- | --- | --- |
| **Age, years, n (%)** |  |  | 0.293 |
| 60–69 | 940 (54.5) | 428 (57.9) |  |
| 70–79 | 520 (30.1) | 206 (27.9) |  |
| ≥ 80 | 265 (15.4) | 105 (14.2) |  |
| **Gender, n (%)** |  |  | 0.142 |
| Male | 854 (49.5) | 342 (46.3) |  |
| Female | 871 (50.5) | 397 (53.7) |  |
| **Race, n (%)** |  |  | 0.786 |
| Non-Hispanic White | 852 (49.4) | 347 (47) |  |
| Non-Hispanic Black | 400 (23.2) | 176 (23.8) |  |
| Mexican American | 149 (8.6) | 63 (8.5) |  |
| Other Hispanic | 169 (9.8) | 79 (10.7) |  |
| Other Races | 155 (9) | 74 (10) |  |
| **Marital status, n (%)** |  |  | 0.761 |
| Married/living with a partner | 1012 (58.7) | 423 (57.2) |  |
| Widowed/divorced/separated | 615 (35.7) | 275 (37.2) |  |
| Never married | 98 (5.7) | 41 (5.5) |  |
| **Education level, n (%)** |  |  | 0.772 |
| Below high school | 407 (23.6) | 176 (23.8) |  |
| High school or equivalent | 395 (22.9) | 178 (24.1) |  |
| Above high school | 923 (53.5) | 385 (52.1) |  |
| **PIR, n (%)** |  |  | 0.727 |
| ≤ 1.3 | 513 (29.7) | 208 (28.1) |  |
| 1.3–3.5 | 658 (38.1) | 289 (39.1) |  |
| > 3.5 | 554 (32.1) | 242 (32.7) |  |
| **Smoking status, n (%)** |  |  | 0.489 |
| Never | 859 (49.8) | 350 (47.4) |  |
| Former | 658 (38.1) | 291 (39.4) |  |
| Current | 208 (12.1) | 98 (13.3) |  |
| **Drinking status, n (%)** |  |  | 0.73 |
| Never | 266 (15.4) | 107 (14.5) |  |
| Former | 477 (27.7) | 214 (29) |  |
| Current | 982 (56.9) | 418 (56.6) |  |
| **PA, n (%)** |  |  | 0.593 |
| Inactive | 572 (33.2) | 241 (32.6) |  |
| Insufficiently active | 286 (16.6) | 135 (18.3) |  |
| Sufficiently active | 867 (50.3) | 363 (49.1) |  |
| **Hypertension, n (%)** |  |  | 0.401 |
| No | 505 (29.3) | 204 (27.6) |  |
| Yes | 1220 (70.7) | 535 (72.4) |  |
| **Stroke, n (%)** |  |  | 0.467 |
| No | 1615 (93.6) | 686 (92.8) |  |
| Yes | 110 (6.4) | 53 (7.2) |  |
| **CHD, n (%)** |  |  | 0.078 |
| No | 1561 (90.5) | 685 (92.7) |  |
| Yes | 164 (9.5) | 54 (7.3) |  |
| **DM, n (%)** |  |  | 0.367 |
| No | 1318 (76.4) | 577 (78.1) |  |
| Yes | 407 (23.6) | 162 (21.9) |  |
| **Depression, n (%)** |  |  | 0.601 |
| No | 1569 (91) | 677 (91.6) |  |
| Yes | 156 (9) | 62 (8.4) |  |
| **p-MCI, n (%)** |  |  | 0.312 |
| No | 1441 (83.5) | 605 (81.9) |  |
| Yes | 284 (16.5) | 134 (18.1) |  |
| **BMI(kg/m<sup>2</sup>), Mean ± SD** | 29.1 ± 6.3 | 29.1 ± 6.2 | 0.943 |
| **ABSI, Mean ± SD** | 0.1 ± 0.0 | 0.1 ± 0.0 | 0.487 |
| **WWI, Mean ± SD** | 11.5 ± 0.7 | 11.5 ± 0.7 | 0.867 |
| **CoI, Mean ± SD** | 1.4 ± 0.1 | 1.4 ± 0.1 | 0.538 |

### Development and evaluation of the nomogram

A clinical prediction model integrating the 10 optimal predictors was developed by synthesizing analytical outcomes, visualized as a nomogram with individualized risk quantification (Fig. 6). Each specific score on the rating scale was quantified by various predictive factors. This scoring tool employs a point allocation system where vertical axis projections translate predictor values into risk scores. Summative risk probability is derived from accumulated points, directly correlating with p-MCI likelihood.

**Fig. 6.**
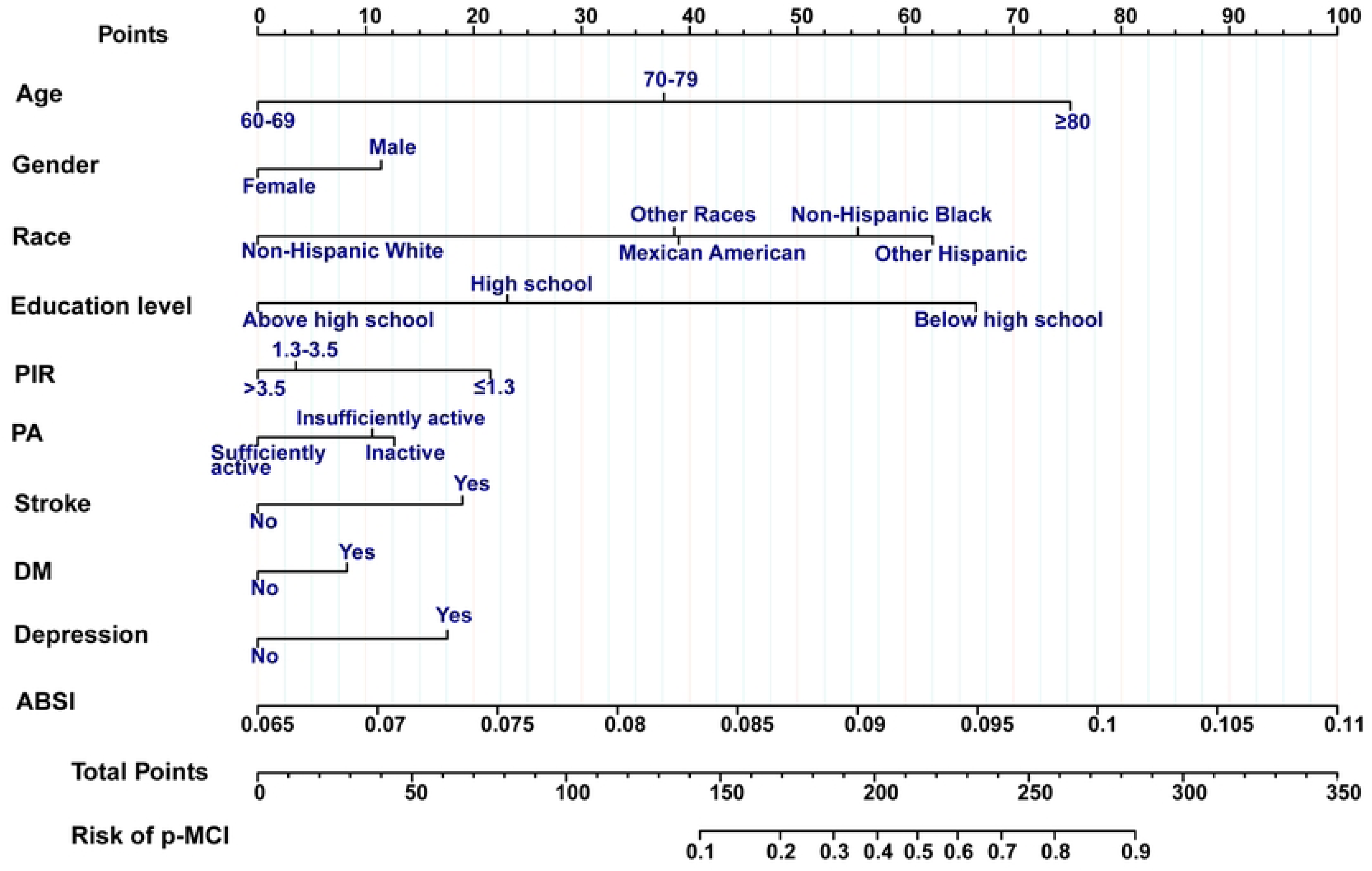
Nomogram prediction model for p-MCI.

The discriminative capability of the model was quantified through ROC analysis, achieving AUC values of 0.861 (95% CI: 0.838–0.884) in the training cohort, 0.826 (95% CI: 0.790–0.861) in the validation cohort, and 0.798 (95% CI: 0.755–0.841) in the external validation cohort (Figs. 7A–C), demonstrating the robust discrimination and prediction capacity. The loess-smoothed calibration curves for both datasets closely resembled straight lines, signifying a good alignment between predicted probabilities and observed outcomes (Figs. 7D–F). Furthermore, the model’s fit was confirmed to be good by the Hosmer-Lemeshow test (*P* = 0.756).

**Fig. 7.**
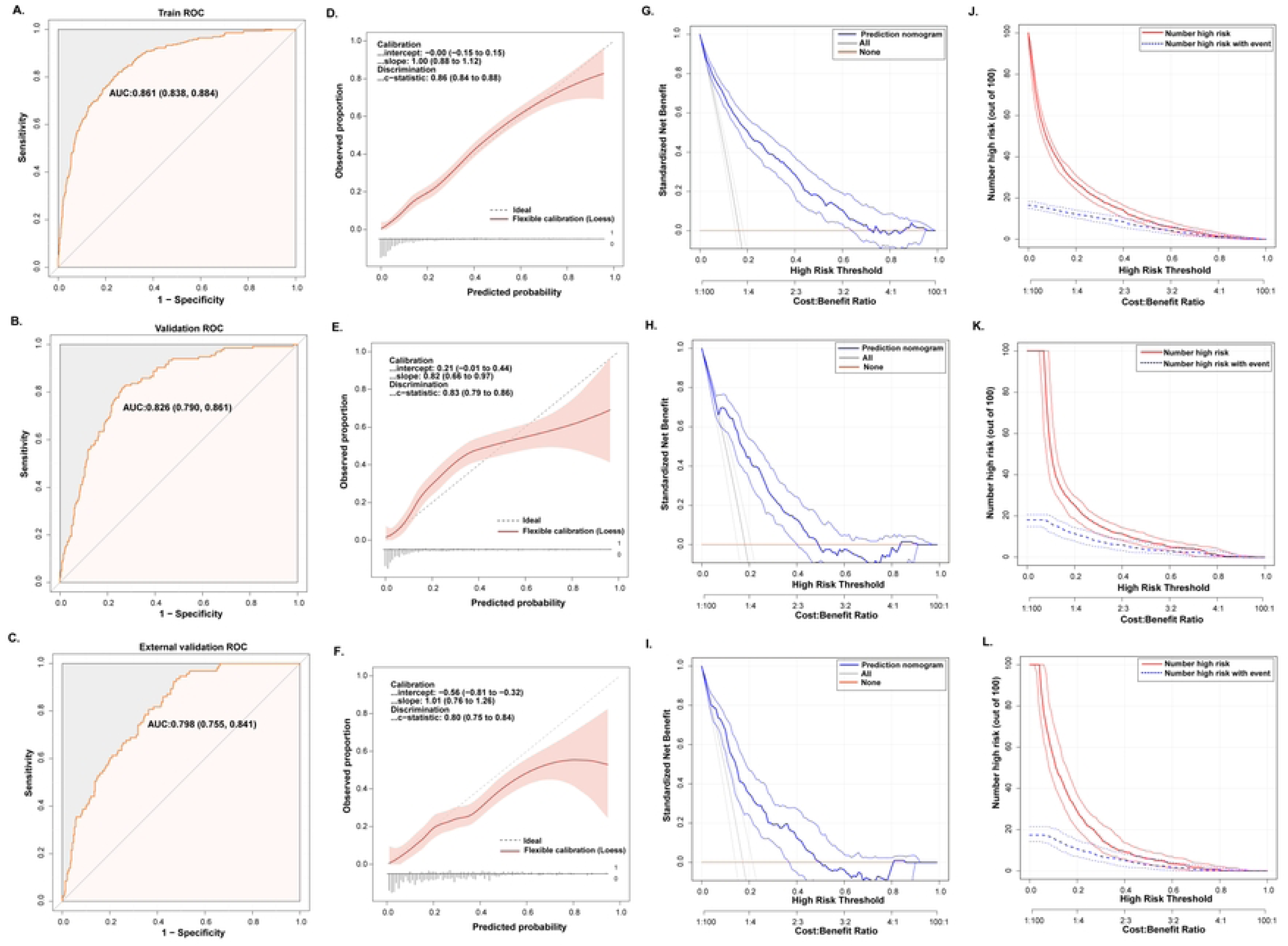
Performance evaluation of nomogram prediction model for p-MCI. **(A)** ROC curve for p-MCI nomogram from a train set. **(B)** ROC curve for p-MCI nomogram from the internal validation set. **(C)** ROC curve for p-MCI nomogram from the external validation set. **(D)** Loess-smoothed calibration curve for p-MCI nomogram from a train set. **(E)** Loess-smoothed calibration curve for p-MCI nomogram from the internal validation set. **(F)** Loess-smoothed calibration curve for p-MCI nomogram from the external validation set. **(G)** DCA for p-MCI nomogram from train set. **(H)** DCA for p-MCI nomogram from the internal validation set. **(I)** DCA for p-MCI nomogram from the external validation set. **(J)** CIC for p-MCI nomogram from a train set. **(K)** CIC for p-MCI nomogram from the internal validation set. **(L)** CIC for p-MCI nomogram from the external validation set.

Clinical applicability was further evaluated using DCA and CIC, confirming good clinical utility. The DCA demonstrated superior net benefit gains compared to universal intervention or non-intervention strategies across clinically relevant risk thresholds in both cohorts (Figs. 7G–I). The CIC analyses revealed close alignment between predicted high-risk cases and confirmed p-MCI diagnoses at risk thresholds exceeding 20% (Figs. 7J–L), validating the precision of the model in risk stratification.

## Discussion

Drawing from NHANES 2011–2014 and CHARLS 2011 wave data, this investigation revealed three key findings in U.S. older adults: (i) Each of three indices related to central obesity (ABSI, WWI, and CoI) demonstrated a positive linear correlation with p-MCI risk; (ii) ten predictors—spanning sociodemographic (age, gender, race, education, and PIR), clinical comorbidities (stroke, DM, and depression), and anthropometric (ABSI) domains—emerged as significant p-MCI risk determinants; (iii) a validated clinical nomogram integrating these predictors achieved robust predictive accuracy, precise calibration, and actionable risk stratification utility. This investigation represents the inaugural effort to evaluate five central adiposity metrics concerning cognitive decline and develop an anthropometry-enhanced predictive tool for elderly populations.

Emerging evidence substantiates significant associations between central adiposity indices and cognitive decline across diverse populations[16,20,47]. Notably, Zhang et al. [20] identified an inverse correlation between ABSI and cognitive functioning in a cohort of the elerly in the U.S. (n = 2,752), where elevated ABSI corresponded to poorer neuropsychological test outcomes. Cross-sectional investigations by Lin et al.[48] and Qiu et al. [49] further demonstrated dose-dependent relationships between WWI elevation and cognitive dysfunction in Chinese and U.S. aging populations. A community-based cohort study also revealed that elevated CoI levels correlated with reduced cognitive performance among older adults in Taiwan [16]. Consistent with prior research, this analysis confirmed ABSI, WWI, and CoI as independent predictors of cognitive impairment after comprehensive covariate adjustment, with findings persisting through multiple imputation sensitivity testing.

Empirical investigations have reported divergent outcomes regarding central adiposity-cognition associations. Guo et al. [50] conducted a cross-sectional analysis of 2,577 rural Chinese adults (35–95 years), revealing non-significant associations between BRI and cognitive deficits. Conversely, Zhang et al. [51] demonstrated significant dose-dependent relationships between elevated BRI and cognitive decline in a U.S. geriatric cohort (≥ 65 years). Discrepancies also extend to WHtR findings: Tang et al. [52] identified an inverse correlation between WHtR and cognitive functioning in the U.S. elderly, whereas Liang et al. [53] found no epidemiological association in their Chinese cohort (n = 10,594; ≥ 65 years). Our study suggests that neither BRI nor WHtR is associated with cognitive impairment. These conflicting outcomes likely stem from methodological heterogeneity across studies, including differences in population stratification (age ranges and geographic distribution), central adiposity metric operationalization, neuropsychological assessment protocols, and covariate adjustment rigor.

Currently, some models incorporating anthropometric indices to predict the risk of cognitive impairment have been reported. Zhang et al. [20] indicated the superior predictive validity of ABSI over conventional metrics (BMI and WC) for assessing cognitive dysfunction risk in the U.S. older populations. Similarly, Qiu et al. [49] documented the enhanced prognostic utility of WWI compared to BMI in cognitive assessments. However, neither study developed a prediction model. In contrast, Chenet al. [54] established a nomogram to estimate the probability of cognitive dysfunction over two- and three-year periods for elderly women with low education in China, which included WHtR and six other features, achieving AUCs of 0.772 (derivation cohort) and 0.803 (internal validation cohort). This investigation employed rigorous multivariable adjustments to examine the associations between central adiposity indices and p-MCI, complemented by stratified sensitivity testing and missing data imputation to verify analytical robustness. Feature selection integrated LASSO regularization with multivariable regression, identifying 10 significant predictors subsequently translated into an intuitive nomogram for point-of-care risk estimation. With an AUC value of 0.8–0.9 considered excellent [55], the model demonstrated strong discriminative capacity (training AUC = 0.861) and reproducible accuracy (internal validation AUC = 0.826, external validation AUC = 0.798). Calibration fidelity was confirmed through smoothed curve alignment (*P* = 0.756, Hosmer–Lemeshow test). Clinical applicability was validated through DCA and CIC, confirming robust net benefit gains and precise risk stratification above 20% probability thresholds.

The visual architecture of the nomogram was used to identify ABSI as a principal determinant of p-MCI. A meta-analytic synthesis established that abdominal adiposity independently increases the risk of cognitive decline, particularly in the elderly [56]. Unlike BMI, which reflects generalized adiposity, central obesity specifically denotes visceral fat accumulation. The original proposal for ABSI—a measure incorporating WC, height, and weight while remaining statistically independent of BMI— was introduced in 2012 to improve the evaluation of central adiposity [28]. Subsequent validation studies demonstrated ABSI’s superiority over other anthropometry indices, including WHtR, in predicting all-cause mortality in Western populations [57]. Beyond mortality, ABSI correlates with cardiovascular morbidity, depression, and carotid atherosclerosis [58–60]. Central obesity is not merely a comorbidity but may directly impair cognitive performance through several mechanisms. Visceral adiposity perpetuates a systemic low-grade inflammatory state, elevating oxidative stress and neuroinflammatory cascades that accelerate gray matter atrophy and microstructural brain alterations [61,62]. Mechanistically, obesity-induced metabolic dysregulation disrupts neural insulin signaling, promotes gut microbiome imbalance (compromising the gut-brain axis), and amplifies pro-inflammatory cytokine production [63]. Chronic adiposity further exacerbates cerebrovascular pathology through oxidative endothelial damage and inflammatory arterial remodeling [64,65].

Epidemiological studies consistently demonstrated a marked age- and education-dependent escalation in MCI prevalence. A meta-analysis of community-based populations among adults aged ≥ 50 years demonstrated that advancing age and lower educational attainment independently predict MCI incidence [2]. Cognitive reserve is understood as the brain’s ability to adapt and compensate, explaining why cognitive decline varies among individuals, often measured by educational attainment [66,67]. Gupta [68] suggested that a larger proportion of Hispanics and Blacks would experience subjective cognitive decline, while Manly et al. [69] documented disproportionate dementia and MCI burdens in these groups—observations concordant with our cohort findings. Our analysis identified elevated neurocognitive risk among males compared to females, potentially mediated by sex-specific cognitive reserve advantages favoring women [70]. Socioeconomic advantage, operationalized through elevated family income, correlated with enhanced cognitive resilience [31]. PA promotes neuroplasticity and neuroprotection, positively affecting patients suffering from cognitive impairment [71]. Stroke and DM significantly amplified cognitive impairment susceptibility and progression severity, aligning with established pathophysiological models [72,73]. Longitudinal evidence substantiated depressive symptom severity as a precursor to MCI development[74]. Depressive pathophysiology initiates neuroinflammatory cascades through pro-inflammatory mediator activation, precipitating cerebral microvascular pathology and hemodynamic insufficiency—both established antecedents to cognitive deterioration [75].

### Advantages and limitations

This investigation demonstrated three principal methodological strengths. Linking NHANES and CHARLS—the nationally representative epidemiological database—enabled robust analysis of population-level associations between central adiposity indices and cognitive outcomes among older adults. The substantial sample size provided adequate statistical power for association analyses and predictive model development or validation. Second, our analytical framework integrated LASSO regularization and multivariable logistic regression to optimize model parsimony and generalizability while minimizing overfitting risks. The final model underwent multidimensional validation through a prespecified protocol assessing predictive accuracy, calibration fidelity, clinical net benefit quantification, and validation consistency. Third, the model-identified features comprised readily accessible variables, offering a pragmatic advantage for the initial screening of geriatric cognitive impairment in resource-constrained community settings. Utilizing the nomogram may enable primary care clinicians to detect MCI cases requiring timely diagnosis and management efficiently.

While this study advances understanding of central adiposity-cognition relationships, three methodological constraints warrant consideration. First, the observational nature of cross-sectional studies inherently limits causal inference between identified predictors and cognitive performance measures. Second, geographic and demographic generalizability remains unestablished given the U.S.-centric sampling frame and Chinese external validation cohort, necessitating more multinational and multiracial external validation. Third, reliance on neuropsychological batteries for case ascertainment introduced diagnostic uncertainty, and future iterations should incorporate neuroimaging and serum biology to refine and validate the model. Finally, residual confounding from unmeasured variables may persist despite rigorous adjustments, highlighting the need to prioritize readily modifiable risk parameters in subsequent models.

## Conclusion

This study revealed significant associations between central adiposity metrics (ABSI, WWI, and CoI) and MCI risk in older adults. The developed nomogram, integrating ABSI with key sociodemographic (age, gender, race, education, and PIR), lifestyle (PA), and comorbidity (stroke, DM, and depression) factors, demonstrated robust predictive capacity for MCI. Model internal and external validation revealed strong performance regarding discrimination, calibration, clinical applicability, and generalization. This prediction model could be a clinically valuable tool for primary care clinicians to implement initial cognitive impairment screening in resource-limited community-based aging populations. Further external validation of our model by prospective international multicenter studies is required before implementation.

## Data Availability

The datasets generated and/or analysed during the current study are available from the NHANES at https://www.cdc.gov/nchs/nhanes, and the CHARLS at http://charls.pku.edu.cn/en.

## Acknowledgements

The authors express gratitude for the accessibility of the NHANES and CHARLS.

## Supporting information

The supplementary data for this article can be found online.

## References

1. Chapman S, Rentería MA, Dworkin JD, Garriga SM, Barker MS, Avila-Rieger J, et al. Association of subjective cognitive decline with progression to dementia in a cognitively unimpaired multiracial community sample. Neurology. 2023;100: e1020– e1027. doi:10.1212/WNL.0000000000201658

2. Bai W, Chen P, Cai H, Zhang Q, Su Z, Cheung T, et al. Worldwide prevalence of mild cognitive impairment among community dwellers aged 50 years and older: a meta-analysis and systematic review of epidemiology studies. Age Ageing. 2022;51: afac173. doi:10.1093/ageing/afac173

3. Anderson ND. State of the science on mild cognitive impairment. J Gerontol Ser B-Psychol Sci Soc Sci. 2020;75: 1359–1360. doi:10.1093/geronb/gbaa040

4. GBD 2019 Dementia Forecasting Collaborators. Estimation of the global prevalence of dementia in 2019 and forecasted prevalence in 2050: an analysis for the global burden of disease study 2019. Lancet Public Health. 2022;7: e105–e125. doi:10.1016/S2468-2667(21)00249-8

5. Matthews KA, Xu W, Gaglioti AH, Holt JB, Croft JB, Mack D, et al. Racial and ethnic estimates of alzheimer’s disease and related dementias in the United States (2015– 2060) in adults aged ≥65 years. Alzheimers Dement. 2019;15: 17–24. doi:10.1016/j.jalz.2018.06.3063

6. Custodio N, Montesinos R, Diaz MM, Herrera-Perez E, Chavez K, Alva-Diaz C, et al. Performance of the rowland universal dementia assessment scale for the detection of mild cognitive impairment and dementia in a diverse cohort of illiterate persons from rural communities in Peru. Front Neurol. 2021;12: 629325. doi:10.3389/fneur.2021.629325

7. Jia L, Quan M, Fu Y, Zhao T, Li Y, Wei C, et al. Dementia in China: epidemiology, clinical management, and research advances. Lancet Neurol. 2020;19: 81–92. doi:10.1016/S1474-4422(19)30290-X

8. Kounnavong S, Vonglokham M, Sayasone S, Savathdy V, Masaki E, Kayano R, et al. Assessment of cognitive function among adults aged ≥ 60 years using the revised hasegawa dementia scale: cross-sectional study, lao people’s democratic republic. Health Res Policy Syst. 2022;20: 121. doi:10.1186/s12961-022-00919-x

9. GBD 2019 Risk Factors Collaborators. Global burden of 87 risk factors in 204 countries and territories, 1990–2019: a systematic analysis for the global burden of disease study 2019. Lancet Lond Engl. 2020;396: 1223–1249. doi:10.1016/S0140-6736(20)30752-2

10. Costache AD, Ignat BE, Grosu C, Mastaleru A, Abdulan I, Oancea A, et al. Inflammatory pathways in overweight and obese persons as a potential mechanism for cognitive impairment and earlier onset alzeihmer’s dementia in the general population: a narrative review. Biomedicines. 2023;11: 3233. doi:10.3390/biomedicines11123233

11. Ma Y, Ajnakina O, Steptoe A, Cadar D. Higher risk of dementia in english older individuals who are overweight or obese. Int J Epidemiol. 2020;49: 1353–1365. doi:10.1093/ije/dyaa099

12. Qu Y, Hu H, Ou Y, Shen X, Xu W, Wang Z, et al. Association of body mass index with risk of cognitive impairment and dementia: a systematic review and meta-analysis of prospective studies. Neurosci Biobehav Rev. 2020;115: 189–198. doi:10.1016/j.neubiorev.2020.05.012

13. Ren Z, Li Y, Li X, Shi H, Zhao H, He M, et al. Associations of body mass index, waist circumference and waist-to-height ratio with cognitive impairment among chinese older adults: based on the CLHLS. J Affect Disord. 2021;295: 463–470. doi:10.1016/j.jad.2021.08.093

14. Jung C-H, Mok J-O. Recent updates on associations among various obesity metrics and cognitive impairment: from body mass index to sarcopenic obesity. J Obes Metab Syndr. 2022;31: 287–295. doi:10.7570/jomes22058

15. Thomas DM, Bredlau C, Bosy-Westphal A, Mueller M, Shen W, Gallagher D, et al. Relationships between body roundness with body fat and visceral adipose tissue emerging from a new geometrical model. Obes Silver Spring Md. 2013;21: 2264– 2271. doi:10.1002/oby.20408

16. Huang S, Chen S, Geng J, Wu D, Li C. Metabolic syndrome and high-obesity-related indices are associated with poor cognitive function in a large taiwanese population study older than 60 years. Nutrients. 2022;14: 1535. doi:10.3390/nu14081535

17. Xu C, Wu X. Association between four anthropometric indices with age-related Macular Degeneration from NHANES 2005–2008. Lipids Health Dis. 2025;24: 11–26. doi:10.1186/s12944-024-02424-2

18. Wang H, Wu S, Pan D, Ning Y, Wang C, Guo J, et al. Risk prediction model of cognitive performance in older people with cardiovascular diseases: a study of the national health and nutrition examination survey database. Front Public Health. 2025;12: 1447366. doi:10.3389/fpubh.2024.1447366

19. Wang J, Wang Y, Li S, Wu B, Feng Q, Qiu W, et al. Waist-to-weight index and cognitive impairment: understanding the link through depression mediation in the NHANES. J Affect Disord. 2024;365: 313–320. doi:10.1016/j.jad.2024.08.067

20. Zhang Y, Zhang P, Yin D. Association between a body shape index and cognitive impairment among US older adults from a cross-sectional survey of the NHANES 2011–2014. Lipids Health Dis. 2024;23: 169. doi:10.1186/s12944-024-02165-2

21. Parker JD, Kruszon-Moran D, Mohadjer LK, Dohrmann SM, Van de Kerckhove W, Clark J, et al. National health and nutrition examination survey: california and los angeles county, estimation methods and analytic considerations, 1999-2006 and 2007-2014. Vital Health Stat 2. 2017; 1–26.

22. Jia W, Wang H, Li C, Shi J, Yong F, Jia H. Association between dietary vitamin B1 intake and cognitive function among older adults: a cross-sectional study. J Transl Med. 2024;22: 165. doi:10.1186/s12967-024-04969-3

23. Li J, Bai Y, Liu Q, Zhang S. Mediation effect of oxidative stress on association between selenium intake and cognition in american adults. Nutrients. 2024;16: 4163. doi:10.3390/nu16234163

24. Wang M, Zeng X, Liu Q, Yang Z, Li J. The association between sleep duration and cognitive function in the U.S. elderly from NHANES 2011–2014: a mediation analysis for inflammatory biomarkers. J Affect Disord. 2025;375: 465–471. doi:10.1016/j.jad.2025.01.154

25. Liu X, Chen J, Meng C, Zhou L, Liu Y. Serum neurofilament light chain and cognition decline in US elderly: A cross-sectional study. Ann Clin Transl Neurol. 2024;11: 17–29. doi:10.1002/acn3.51929

26. Liang K, Zhang X. Association between life’s essential 8 and cognitive function: insights from NHANES 2011–2014. Front Aging Neurosci. 2024;16: 1386498. doi:10.3389/fnagi.2024.1386498

27. Smagula SF, Zhang G, Gujral S, Covassin N, Li J, Taylor WD, et al. Association of 24-hour activity pattern phenotypes with depression symptoms and cognitive performance in aging. JAMA Psychiatry. 2022;79: 1023. doi:10.1001/jamapsychiatry.2022.2573

28. Krakauer NY, Krakauer JC. A new body shape index predicts mortality hazard independently of body mass index. PLoS ONE. 2012;7: e39504. doi:10.1371/journal.pone.0039504

29. Valdez R. A simple model-based index of abdominal adiposity. J Clin Epidemiol. 1991;44: 955–956. doi:10.1016/0895-4356(91)90059-I

30. Park Y, Kim NH, Kwon TY, Kim SG. A novel adiposity index as an integrated predictor of cardiometabolic disease morbidity and mortality. Sci Rep. 2018;8: 16753. doi:10.1038/s41598-018-35073-4

31. Iskandar M, Martindale J, Bynum JPW, Davis MA. Association between Family Household Income and Cognitive Resilience among Older US Adults: A Cross-Sectional Study. J Prev Alzheimers Dis. 2024;5: 1406–1409. doi:10.14283/jpad.2024.97

32. Huang Y, Liu X, Lin C, Chen X, Li Y, Huang Y, et al. Association between the dietary index for gut microbiota and diabetes: the mediating role of phenotypic age and body mass index. Front Nutr. 2025;12: 1519346. doi:10.3389/fnut.2025.1519346

33. Ge J, Peng W, Lu J. Predictive value of life’s crucial 9 for cardiovascular and all-cause mortality: a prospective cohort study from the NHANES 2007 to 2018. J Am Heart Assoc. 2024;13: e036669. doi:10.1161/JAHA.124.036669

34. Liu J, Huang S. Dietary index for gut microbiota is associated with stroke among US adults. Food Funct. 2025; 10.1039.D4FO04649H. doi:10.1039/D4FO04649H

35. Wei X, Min Y, Xiang Z, Zeng Y, Wang J, Liu L. Joint association of physical activity and dietary quality with survival among US cancer survivors: a population-based cohort study. Int J Surg Lond Engl. 2024;110: 5585–5594. doi:10.1097/JS9.0000000000001636

36. Tang H, Zhang X, Luo N, Huang J, Zhu Y. Association of dietary live microbes and nondietary prebiotic/probiotic intake with cognitive function in older adults: evidence from NHANES. Fielding R, editor. J Gerontol Ser -Biol Sci Med Sci. 2024;79: glad175. doi:10.1093/gerona/glad175

37. Zhou J, Li Y, Zhu L, Yue R. Association between frailty index and cognitive dysfunction in older adults: insights from the 2011–2014 NHANES data. Front Aging Neurosci. 2024;16: 1458542. doi:10.3389/fnagi.2024.1458542

38. Zhang X, Yang Q, Huang J, Lin H, Luo N, Tang H. Association of the newly proposed dietary index for gut microbiota and depression: the mediation effect of phenotypic age and body mass index. Eur Arch Psychiatry Clin Neurosci. 2024 [cited 22 Oct 2024]. doi:10.1007/s00406-024-01912-x

39. Zhao Y, Hu Y, Smith JP, Strauss J, Yang G. Cohort Profile: The China Health and Retirement Longitudinal Study (CHARLS). Int J Epidemiol. 2012;43: 61. doi:10.1093/ije/dys203

40. Li J, Sun J, Zhang Y, Zhang B, Zhou L. Association between weight-adjusted-waist index and cognitive decline in US elderly participants. Front Nutr. 2024;11: 1390282. doi:10.3389/fnut.2024.1390282

41. Cheng Y, Fang Z, Zhang X, Wen Y, Lu J, He S, et al. Association between triglyceride glucose-body mass index and cardiovascular outcomes in patients undergoing percutaneous coronary intervention: a retrospective study. Cardiovasc Diabetol. 2023;22: 75. doi:10.1186/s12933-023-01794-8

42. Hu J, Wang Y, Tong X, Yang T. When to consider logistic LASSO regression in multivariate analysis? Eur J Surg Oncol. 2021;47: 2206. doi:10.1016/j.ejso.2021.04.011

43. Li Z, Shen Q, Wang J, Tuo J, Tan Y, Li H, et al. Prediagnostic plasma metabolite concentrations and liver cancer risk: a population-based study of chinese men. Ebiomedicine. 2024;100: 104990. doi:10.1016/j.ebiom.2024.104990

44. Wang L, Ren J, Chen J, Gao R, Bai B, An H, et al. Lifestyle choices mediate the association between educational attainment and BMI in older adults in China: a cross-sectional study. Front Public Health. 2022;10: 1000953. doi:10.3389/fpubh.2022.1000953

45. Yan J, Liu Y, Yu J, Liao L, Wang H. Establishment and validation of a nomogram for suicidality in chinese secondary school students. J Affect Disord. 2023;330: 148–157. doi:10.1016/j.jad.2023.02.062

46. Liu X, Yu T, Qi J, Lv R, Wang Q. Factors predicting neuronal surface antibodies in the elderly with new-onset and unknown seizures. Ann Clin Transl Neurol. 2022;9: 1039. doi:10.1002/acn3.51597

47. Zhao C, Xu X, Hao C. Evidence from NHANES 2011–2014: a correlation between the weight-adjusted-waist index and cognitive abilities in the United States. Front Aging Neurosci. 2025;17: 1480609. doi:10.3389/fnagi.2025.1480609

48. Lin J, Shen H, Yang W, Zhang G, Sun J, Shen W, et al. Association between weight-adjusted waist index and cognitive impairment in chinese older men: a 7-year longitudinal study. Front Aging Neurosci. 2025;17: 1510781. doi:10.3389/fnagi.2025.1510781

49. Qiu X, Kuang J, Huang Y, Wei C, Zheng X. The association between weight-adjusted-waist index (WWI) and cognitive function in older adults: a cross-sectional NHANES 2011–2014 study. BMC Public Health. 2024;24: 2152. doi:10.1186/s12889-024-19332-w

50. Guo D, Li T, Yang Q, Yang C, Yang Y, Liu F, et al. Relationship between body roundness index and cognitive impairment in middle-aged and older adults: a population-based cross-sectional study. Front Aging Neurosci. 2025;17: 1522989. doi:10.3389/fnagi.2025.1522989

51. Zhang F, Ning Z, Wang C. Body roundness index and cognitive function in older adults: a nationwide perspective. Front Aging Neurosci. 2024;16: 1466464. doi:10.3389/fnagi.2024.1466464

52. Tang H, Li Q, Du C. The association between waist-to-height ratio and cognitive function in older adults. Nutr Neurosci. 2024;27: 1405–1412. doi:10.1080/1028415X.2024.2339729

53. Liang Z, Jin W, Huang L, Chen H. Body mass index, waist circumference, hip circumference, abdominal volume index, and cognitive function in older chinese people: a nationwide study. BMC Geriatr. 2024;24: 925. doi:10.1186/s12877-024-05521-0

54. Chen Z, Du J, Song Q, Yang J, Wu Y. A prediction model of cognitive impairment risk in elderly illiterate chinese women. Front Aging Neurosci. 2023;15: 1148071. doi:10.3389/fnagi.2023.1148071

55. Cooper JD, Han SYS, Tomasik J, Ozcan S, Rustogi N, Beveren NJM van, et al. Multimodel inference for biomarker development: an application to schizophrenia. Transl Psychiatry. 2019;9: 83. doi:10.1038/s41398-019-0419-4

56. Tang X, Zhao W, Lu M, Zhang X, Zhang P, Xin Z, et al. Relationship between central obesity and the incidence of cognitive impairment and dementia from cohort studies involving 5,060,687 participants. Neurosci Biobehav Rev. 2021;130: 301–313. doi:10.1016/j.neubiorev.2021.08.028

57. Krakauer NY, Krakauer JC. Dynamic association of mortality hazard with body shape. PLOS One. 2014;9: e88793. doi:10.1371/journal.pone.0088793

58. Bozorgmanesh M, Sardarinia M, Hajsheikholeslami F, Azizi F, Hadaegh F. CVD-predictive performances of “a body shape index” versus simple anthropometric measures: tehran lipid and glucose study. Eur J Nutr. 2016;55: 147–157. doi:10.1007/s00394-015-0833-1

59. Ma X, Chen L, Hu W, He L. Association between a body shape index and subclinical carotid atherosclerosis in population free of cardiovascular and cerebrovascular diseases. J Atheroscler Thromb. 2021;29: 1140. doi:10.5551/jat.62988

60. Zhang Z, Ruan X, Ma W. The association between a body shape index and depressive symptoms: a cross-sectional study using NHANES data (2011–2018). Front Nutr. 2025;11: 1510218. doi:10.3389/fnut.2024.1510218

61. Naomi R, Teoh SH, Embong H, Balan SS, Othman F, Bahari H, et al. The role of oxidative stress and inflammation in obesity and its impact on cognitive impairments—a narrative review. Antioxidants. 2023;12: 1071. doi:10.3390/antiox12051071

62. Salas-Venegas V, Flores-Torres RP, Rodríguez-Cortés YM, Rodríguez-Retana D, Ramírez-Carreto RJ, Concepción-Carrillo LE, et al. The obese brain: mechanisms of systemic and local inflammation, and interventions to reverse the cognitive deficit. Front Integr Neurosci. 2022;16: 798995. doi:10.3389/fnint.2022.798995

63. Zhang Q, Jin K, Chen B, Liu R, Cheng S, Zhang Y, et al. Overnutrition induced cognitive impairment: insulin resistance, gut-brain axis, and neuroinflammation. Front Neurosci. 2022;16: 884579. doi:10.3389/fnins.2022.884579

64. Olsthoorn L, Vreeken D, Kiliaan AJ. Gut microbiome, inflammation, and cerebrovascular function: link between obesity and cognition. Front Neurosci. 2021;15: 761456. doi:10.3389/fnins.2021.761456

65. Taïlé J, Bringart M, Planesse C, Patché J, Rondeau P, Veeren B, et al. Antioxidant polyphenols of antirhea borbonica medicinal plant and caffeic acid reduce cerebrovascular, inflammatory and metabolic disorders aggravated by high-fat diet-induced obesity in a mouse model of stroke. Antioxidants. 2022;11: 858. doi:10.3390/antiox11050858

66. Stern Y, Arenaza-Urquijo EM, Bartrés-Faz D. Whitepaper: defining and investigating cognitive reserve, brain reserve and brain maintenance. Alzheimers Dement. 2020;16: 1305. doi:10.1016/j.jalz.2018.07.219

67. Vonk JM, Ghaznawi R, Zwartbol MH, Stern Y, Geerlings MI. The role of cognitive and brain reserve in memory decline and atrophy rate in mid and late-life: the SMART-MR study. Cortex. 2022;148: 204. doi:10.1016/j.cortex.2021.11.022

68. Gupta S. Racial and ethnic disparities in subjective cognitive decline: a closer look, united states, 2015–2018. BMC Public Health. 2021;21: 1173. doi:10.1186/s12889-021-11068-1

69. Manly JJ, Jones RN, Langa KM, Ryan LH, Levine DA, McCammon R, et al. Estimating the prevalence of dementia and mild cognitive impairment in the US: the 2016 health and retirement study harmonized cognitive assessment protocol project. JAMA Neurol. 2022;79: 1242. doi:10.1001/jamaneurol.2022.3543

70. Levine DA, Gross AL, Briceño EM, Tilton N, Giordani BJ, Sussman JB, et al. Sex differences in cognitive decline among US adults. JAMA Netw Open. 2021;4: e210169. doi:10.1001/jamanetworkopen.2021.0169

71. Farì G, Lunetti P, Pignatelli G, Raele MV, Cera A, Mintrone G, et al. The effect of physical exercise on cognitive impairment in neurodegenerative disease: from pathophysiology to clinical and rehabilitative aspects. Int J Mol Sci. 2021;22: 11632. doi:10.3390/ijms222111632

72. Ehtewish H, Arredouani A, El-Agnaf O. Diagnostic, prognostic, and mechanistic biomarkers of diabetes mellitus-associated cognitive decline. Int J Mol Sci. 2022;23: 6144. doi:10.3390/ijms23116144

73. Lo JW, Crawford JD, Desmond DW, Bae H, Lim J, Godefroy O, et al. Long-term cognitive decline after stroke: an individual participant data meta-analysis. Stroke. 2022;53: 1318–1327. doi:10.1161/STROKEAHA.121.035796

74. Guo Y, Pai M, Xue B, Lu W. Bidirectional association between depressive symptoms and mild cognitive impairment over 20 years: Evidence from the Health and Retirement Study in the United States. J Affect Disord. 2023;338: 449–458. doi:10.1016/j.jad.2023.06.046

75. Hakim A. Perspectives on the complex links between depression and dementia. Front Aging Neurosci. 2022;14: 821866. doi:10.3389/fnagi.2022.821866

